# Neoadjuvant Chemotherapy plus Surgery versus Direct Surgery in Senile Patients with Gastric Cancer: Single-Center Retrospective Cohort Study

**DOI:** 10.1101/2023.04.08.23288188

**Authors:** Birendra Kumar Sah, Zhenjia Yu, Jian Li, Sheng Lu, Yanan Zheng, Zhenglun Zhu, Chen Li, Min Yan, Zhenggang Zhu

## Abstract

**Background:** Despite the lack of decisive research advocating neoadjuvant chemotherapy there is a broad consensus that it is beneficial for gastric cancer in terms of survival. However, there is no comparative research on whether it is similarly helpful in senile patients with the age above 75 years old. Here we compared the survival rate between neoadjuvant plus surgery with Direct Surgery.

**Methods:** We analyzed 79 patients with locally advanced gastric cancer who were preoperatively suspicious of serosa positive or beyond (cT4a or cT4b); or extensive lymph node involvement (cN3). Postoperative complications and overall survival rate were compared between the patients who underwent neoadjuvant chemotherapy (NAC) plus surgery and the patients who had direct surgery.

**Results:** A total of 15 (19%) patients underwent neoadjuvant chemotherapy and 64 (81%) patients had direct surgery. The median follow-up time was 34 months (range of 24-60 months). While the median survival time was not reached in the direct surgery group, the median survival time for the NAC plus Surgery Group was 37 months. Two years of overall survival (OS) for the patients in the NAC plus Surgery group and direct surgery group were 53.3% and 70.3% respectively. There was no statistical difference between the two groups (p>0.05) in overall postoperative complication and length of postoperative stay.

**Conclusions:** Reduced does of neoadjuvant chemotherapy was feasible in senile patients. There was no difference in survival rate between the patients who had neoadjuvant plus surgery compared to those who had direct surgery. While this result contradicts the previous assumption that neoadjuvant chemotherapy is beneficial for late-stage gastric cancer patients, a well-controlled prospective study is mandatory for a better understanding of whether neoadjuvant chemotherapy is beneficial to senile patients too.

## Background

Neoadjuvant chemotherapy was a highly debated topic for the last two decades, especially between Japan and other Western countries and despite multiple RCTs worldwide (1-4), confusion remains in the field (5, 6). Nevertheless, there is a broad consensus that Neoadjuvant chemotherapy may benefit advanced gastric cancer and it may increase the overall survival rate (7-10). However, most of the past RCTs have not enrolled elderly patients, especially above 75 years of age (11). We carefully conducted a retrospective study to understand whether the Neoadjuvant chemotherapy is similarly beneficial for this group of patients so that we can have a basis to conduct a prospective study to further address this issue. In this work, we compared the overall survival rate between the elderly patients who had received Neoadjuvant chemotherapy plus surgery with the patients who had direct surgery. To achieve as many concrete results from a retrospective study we partially selected only the patients with much-advanced stage, as neoadjuvant chemotherapy is much more controversial in patients with stage II or below, especially in East Asian countries.

## Methods

### Study design

The primary inclusion criterion was that the elderly patients above 75 years old who had undergone surgical treatment for gastric cancer and completed follow-up for at least 24 months after the treatment. We only included pathologically confirmed advanced staged gastric cancer patients with clinical staging T4A or T4B or N3, who were preoperatively assessed by computed tomography (CT). The patients with distant metastases were not included. All patients had a performance status of Eastern Cooperative Oncology Group (ECOG) ≤ 2 scores. All the patients were treated between January 2018 and December 2021 at Ruijin Hospital, Shanghai Jiaotong University School of Medicine.

In this cohort, the patients were only included if they met all the inclusion criteria and we only collected the data of patients who were included in this study.

### Treatment

Patients received chemotherapy for two months before surgery, generally, 3 cycles of chemotherapy were prescribed and the dose of chemotherapy was decreased to 20% less than the standard dose. Standard gastrectomy with curative intent was the principal surgical procedure. It involves resection of at least two-thirds of the stomach with D2 lymph node dissection. Postoperative morbidity and mortality were recorded according to the Clavien-Dindo grading(12).

Overall survival (OS) time in this study is the time from the date of surgery to death from any cause. The data for Relapse-free survival (RFS) was not available accurately, thus not analyzed in this study.

### Statistical analysis

The statistical analysis was performed with Statistical Package for Social Science (SPSS) version 22.0 for Windows (SPSS, Inc., Chicago, Illinois). Nonparametric methods were used to analyze data with an abnormal distribution. The continuous data are expressed as the median and range, the Mann-Whitney U test was used for continuous data. The chi-square test and Fisher’s exact test were used to compare the differences between the two groups as appropriate. A p-value of less than 0.05 was considered statistically significant. Survival data were presented as the length of overall survival (OS), in months. A Kaplan-Meier plot was created for survival analysis, Log Rank method was used to compare the OS. Cox regression analysis was used to identify the risk factors for OS.

## Results

The general characteristics of the patients were described in Table 1; the median age was 77 years old in NAC plus Surgery group and 79 years old in the Direct Surgery group. Proximal tumors were more dominant in NAC plus Surgery group than those in the Direct Surgery group (p<0.05). Significantly more patients underwent total gastrectomy in NAC plus Surgery group compared to those in the Direct Surgery group (p<0.05). Approximately one-third of all patients received postoperative chemotherapy, there was no difference in postoperative chemotherapy between the two groups (p>0.05). Altogether 15 patients received neoadjuvant chemotherapy, 60 percent of them had SOX, and 87 percent of patients had three cycles of chemotherapy before the surgery (Table 2). We found that the chemotherapy dose was reduced in these patients by 20-30 percent less than that of the standard dose. Details on the adverse effects of the chemotherapy were not described due to the lack of actual data due to the retrospective nature of the current study. Details on postoperative pathology were described in Table 3.

**Table 1.** Demographic Data

| Parameter |  | NAC plus Surgery | Direct Surgery | P Value |
| --- | --- | --- | --- | --- |
| Sex | Male | 12 (80.0) | 41 (64.1) | 0.237 |
|  | Female | 3 (20.0) | 23 (35.9) |  |
| Age (years) | Median | 77 | 79 | 0.217 |
|  | Range | 76-85 | 76-89 |  |
| Body mass index | Median | 22.49 | 21.66 | 0.336 |
| Site of tumor | Proximal | 8 (53.3) | 13(20.3) | 0.041* |
|  | Body | 4 (26.7) | 22 (34.4) |  |
|  | Distal | 3 (20.0) | 29 (45.3) |  |
| cTNM stage | III | 14(93.3) | 56(87.5) | 0.456 |
|  | IVA | 1(6.7) | 8(12.5) |  |
| R0 Resection | Yes | 14 (93.3) | 59 (92.2) | 0.681 |
|  | No | 1 (6.7) | 5(7.8) |  |
| Type of Gastrectomy | Partial | 4 (26.7) | 46(71.9) | 0.002 |
|  | Total | 11 (73.3) | 18(28.1) |  |
| Postop Chemo | Yes | 5(33.3) | 20(31.3) | 0.899* |
|  | No | 9(60.0) | 41(64.1) |  |
|  | Unknown | 1(6.7) | 3(4.7) |  |

\*Fisher's Exact Test

**Table 2.**
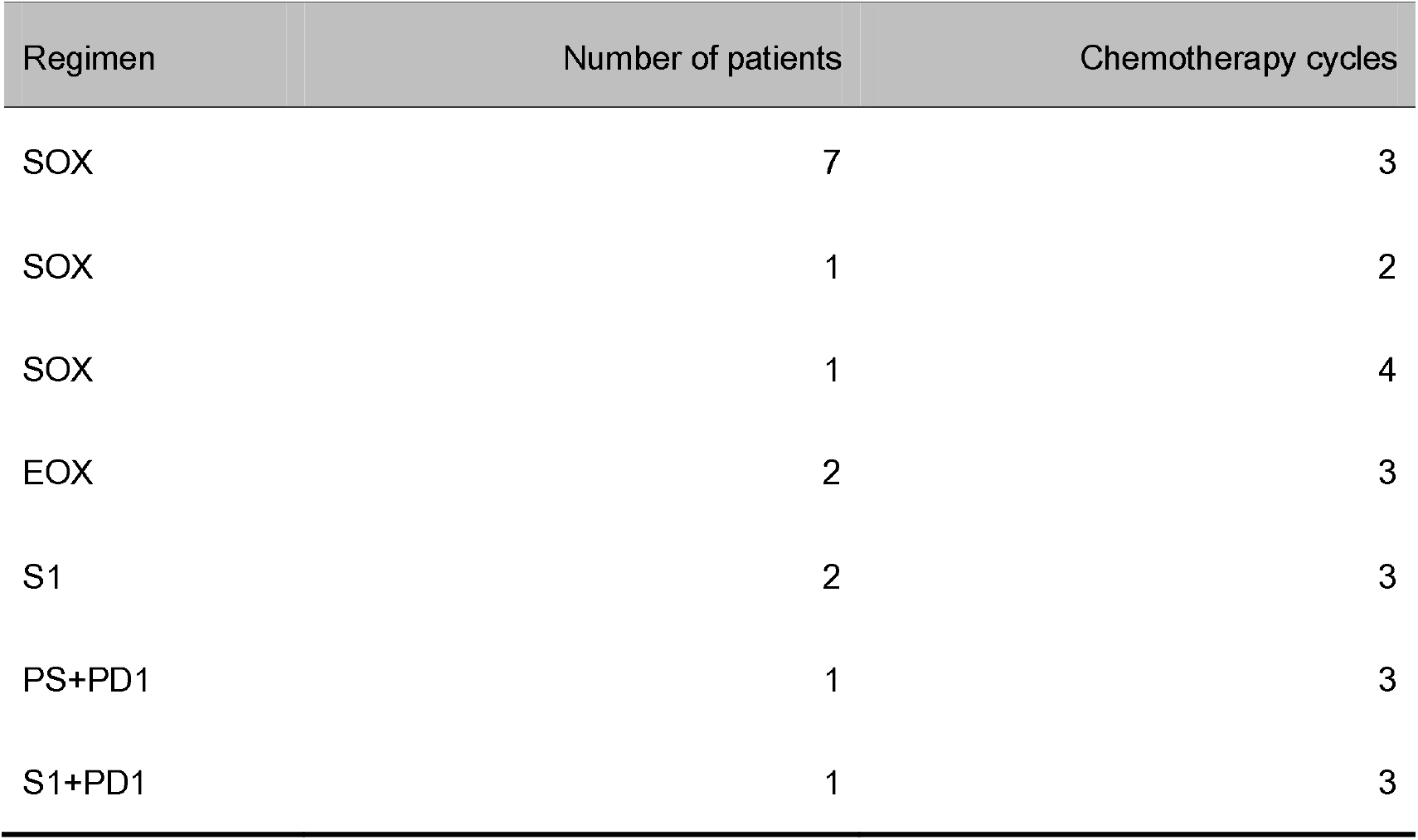
Preoperative Chemotherapy

**Table 3.** Postoperative Pathology

| Parameter | Stage | NAC plus Surgery | Direct Surgery |
| --- | --- | --- | --- |
| ypTNM | pCR | 1 | NA |
|  | I | 2 | NA |
|  | II | 4 | NA |
|  | III | 8 | NA |
| pTNM | IA | NA | 2(3.1) |
|  | IIA | NA | 4(6.3) |
|  | IIB | NA | 11(17.2) |
|  | IIIA | NA | 23(35.9) |
|  | IIIB | NA | 14(21.9) |
|  | IIIC | NA | 10(15.6) |

There was no significant difference (p=0.495) in overall postoperative complications between the two groups, 40.0 % in NAC plus Surgery group versus 35.9% in the Direct Surgery group (Table 4). Similarly, there was no difference in surgical complications between the two groups (p=0.969). All 79 patients had a timely follow-up for two years and above, the median follow-up time was 34 months (range of 24-60 months). There was no statistically significant difference in two years of overall survival (OS) between the patients in the NAC group (53.3%) and the direct surgery group (70.3%). While the median survival time was not reached in the direct surgery group, the median survival time for the NAC Group is 37 months, there was no significant difference between the two groups (p=0.294). A Kaplan-Meier plot for OS is provided in Fig.1. Cox Regression analysis found that the type of gastrectomy was an independent risk factor for overall survival (P=0.005, HR 1.828, 95% CI 1.203-2.776); the patient who underwent total gastrectomy had shorter survival time than those who underwent partial gastrectomy.NAC plus Surgery or Direct surgery was not a risk factor for overall survival.

**Table 4.** Postoperative Complications

| Complication | NAC plus Surgery | Direct Surgery |
| --- | --- | --- |
| Overall complications | 6(40.0) | 23(35.9) |
| Grade 0- I | 9(60.0) | 41 ( 64.1 ) |
| Grade II | 4 ( 26.7 ) | 15 ( 23.4 ) |
| Grade IIIa | 2 ( 13.3 ) | 3 ( 4.7 ) |
| Grade IVa | 0 | 4 ( 6.2 ) |
| Grade V | 0 | 1 ( 1.6 ) |
| Surgical complication | 5(33.3) | 21(32.8) |
| Intra-abdominal hemorrhage | 1(6.7) | 1(1.6) |
| Reoperation | 0 | 3 ( 4.7 ) |
| Anastomotic leakage | 1 ( 6.7 ) | 5(7.8) |
| Duodenal stump leak | 0 | 2(3.2) |
| Anastomotic bleeding | 1(6.7) | 0 |
| Pulmonary Infection | 2(13.3) | 8(12.5) |
| Abdominal Infection | 4(26.7) | 16(25.0) |
| Urinary tract infection | 0 | 1 |
| Central line infection | 1(6.7) |  |
| Bloodstream infection | 1(6.7) | 2(3.2) |
| Wound infection | 1(6.7) | 1(1.6) |
| Pancreatic fistula | 1 ( 6.7 ) | 2(3.1) |
| Death | 0 | 1 |

**Fig. 1.**
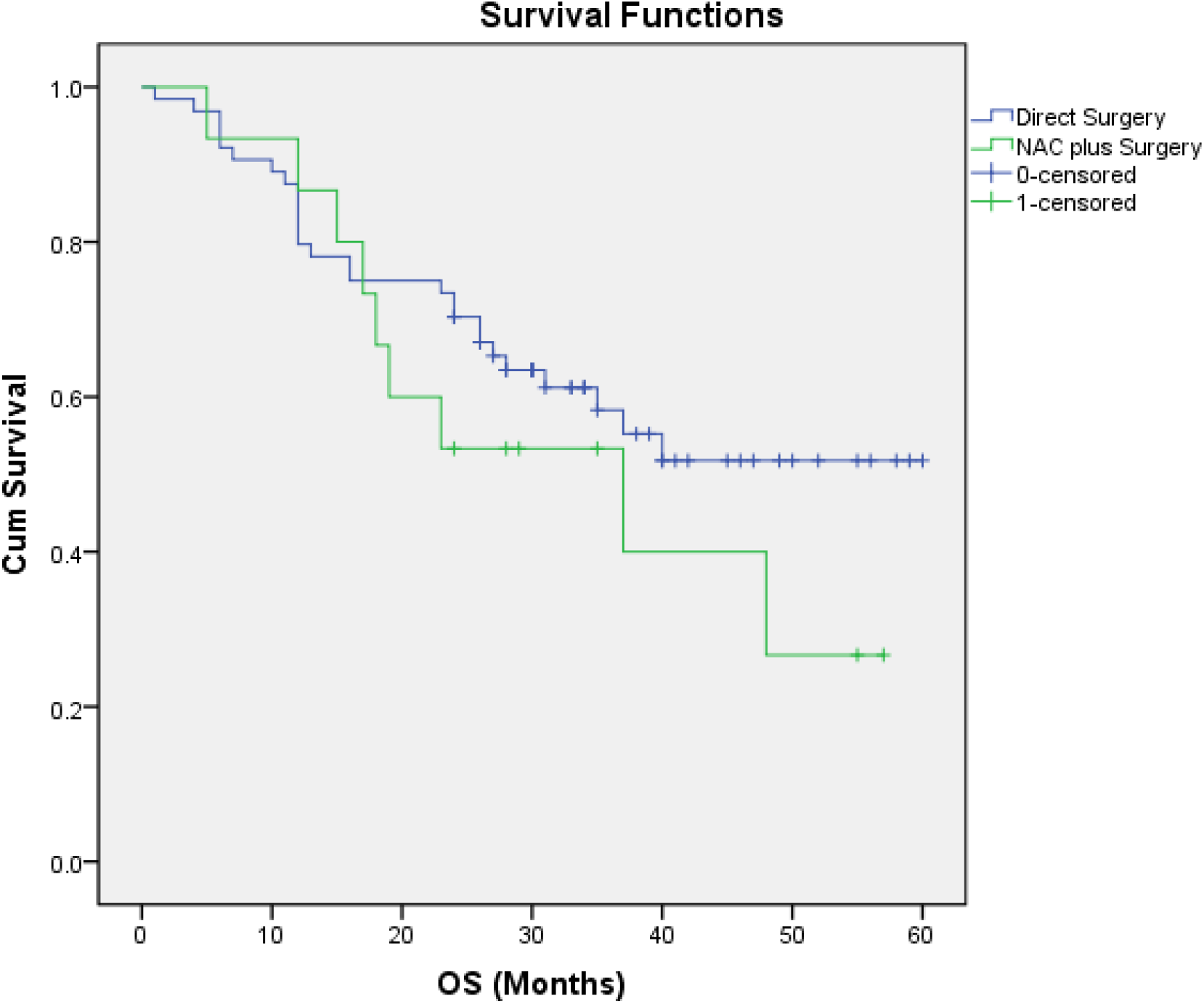
Kaplan-Meier (K-M) Plot for OS

## Discussion

Neoadjuvant chemotherapy has been extensively researched for gastric cancer patients, recently from Germany too (13), and many RCTs advocated for its beneficial effects, including in Japan and China (4,14). However, most of those studies conducted prospective research excluding a group of senile patients, especially those above 75 years (7), perhaps assuming that those patients might not tolerate preoperative chemotherapy. But we noticed there were a substantial number of patients who underwent surgical treatment for advanced-stage gastric cancer in our center. So there was a scientific question that whether major surgery like radical gastrectomy was better tolerated than neoadjuvant chemotherapy in senile patients. In general, these major surgeries are only performed if the overall organ function is satisfactory for surgical insult. And those criteria are almost similar for evaluating a patient for neoadjuvant chemotherapy. Therefore, we wanted to conduct a prospective study to compare all aspects of results between the patients who undergo neoadjuvant chemotherapy plus surgery with the patients who go for direct surgery. As we did not find such studies in past literature, we conducted this retrospective analysis for a scientific basis to conduct a prospective study.

We intentionally only included relatively late-stage locally advanced gastric cancer patients in this study, so that the result would be more acceptable for the clinician in the region. Despite a clear suggestion in NCCN guidelines suggesting any patients with locally advanced gastric cancer should receive preoperative treatment or neoadjuvant chemotherapy before surgery, neoadjuvant chemotherapy is not well accepted, especially in East Asian countries, and the neoadjuvant chemotherapy is generally administered in patients with relatively late stage (15,16). Therefore, we selected the group of serosa-invaded patients or the patients with extensively enlarged regional lymph nodes.

In general, there is a trend that any prospective study generally enrolls patients below 70 or 75 years old. Thus, the clinical decision for these patients is highly heterogeneous in the different centers and much more dependent on the patient or their family, with the probably prejudiced assumption that these senile patients might not tolerate neoadjuvant chemotherapy. The result shows that these patients tolerated major surgery and the neoadjuvant chemotherapy plus surgery too. This finding incites further research in this field and somehow establishes a thin scientific rationale for more extensive exploration of this particular group of patients.

We believe that the two years overall survival analysis is not enough for reaching any conclusion, however, the result itself is quite interesting that the overall survival rate was much lower in the group of patients who had neoadjuvant chemotherapy plus surgery, this result contradicts the results in previously published articles. It might be related to the higher number of total gastrectomies in the NAC plus Surgery group, as the Cox Regression analysis found that the type of gastrectomy was an independent risk factor for overall survival in this study. Besides, we noticed that there were 17 patients in the Direct Surgery group with pathological diagnosis pTNM below stage III. A similar over-diagnosis was earlier reported by Japanese researchers, more than 10 percent of patients with the clinical diagnosis of T3 and T4 were eventually diagnosed with pathological stage I (17). We therefore further calculated the overall survival rate excluding those patients, similarly, there was no significant difference (p<0.05) between the two groups Fig.2. And we confirmed that these patients were due to misdiagnosis before surgery. Nevertheless, if the prospective study similarly confirms the results of this study then it will further validate the gut feeling of the surgeons who were reluctant to administer neoadjuvant chemotherapy, especially to these senile patients. Therefore, despite a less number of patients in this cohort and a shorter period of follow-up time we still decided to publish these findings as we consider this would further stimulate more prospective research.

**Fig. 2.**
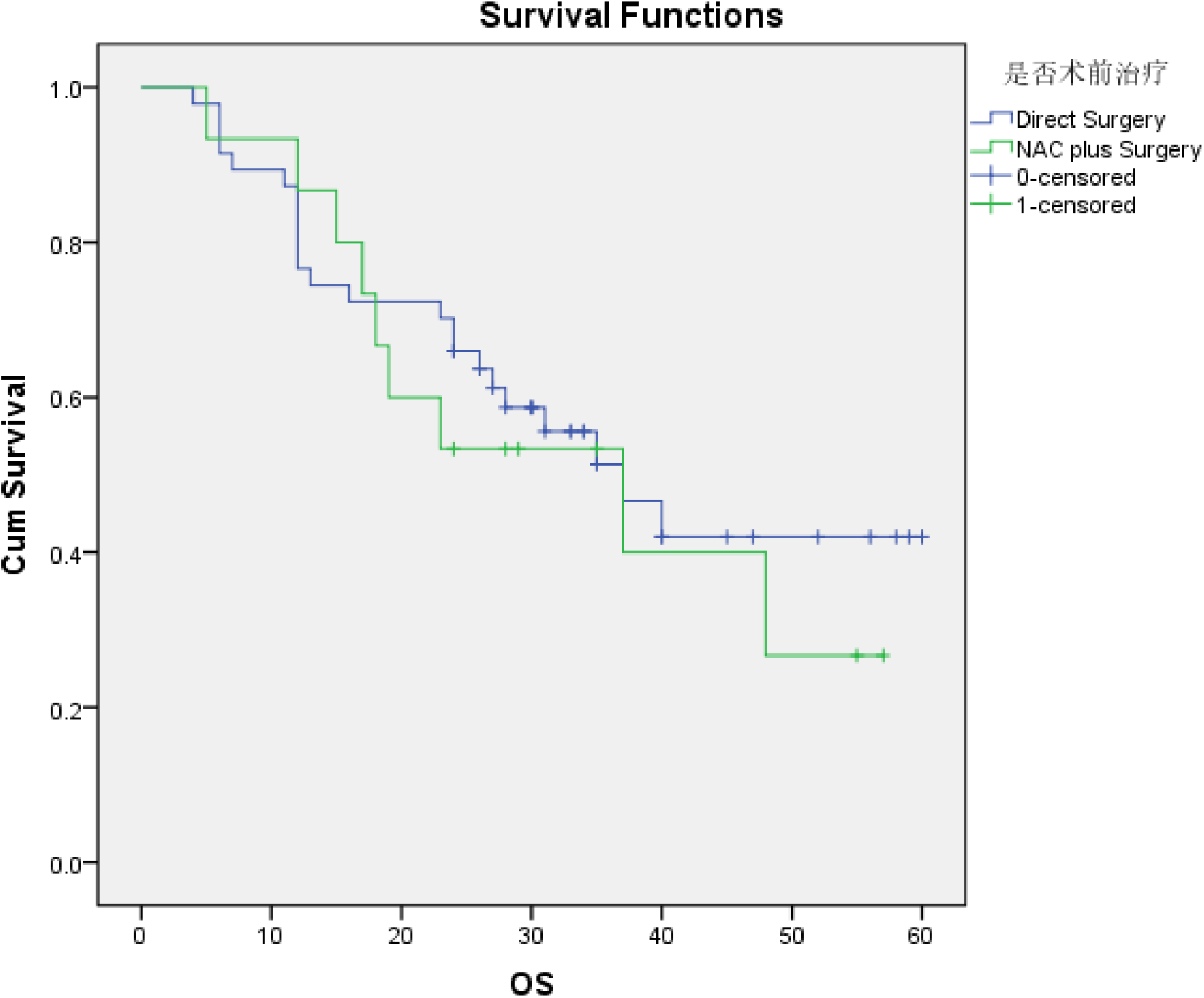
Kaplan-Meier (K-M) Plot for OS (only pTNM III in Direct Surgery group)

We recognize a few major limitations of this study, the number of patients was relatively less and there was a significant difference in an important factor (types of gastrectomy) between the two groups which might have affected the result. Besides, it was difficult to understand whether all senile patients can receive neoadjuvant chemotherapy or surgery. Therefore the result of this study should be interpreted accordingly.

## Conclusion

Reduced doses of neoadjuvant chemotherapy were feasible in senile patients, there was no difference in survival rate between the patients who had neoadjuvant plus surgery compared to those who had direct surgery. Since this result contradicts the previous assumption that neoadjuvant chemotherapy is beneficial for late-stage gastric cancer patients, a well-controlled prospective study is mandatory for a better understanding of whether neoadjuvant chemotherapy is beneficial to senile patients too.

## Data Availability

The datasets used and/or analyzed during the current study are available from the corresponding author on reasonable request.

## Declaration

### Ethics approval and consent to participate

The Ethics Committee of Ruijin Hospital approved this study and waived the individual informed consent due to the retrospective analysis. The study was carried out in conformity with the Declaration of Helsinki (as revised in 2013)

### Consent for publication

The manuscript does not contain any individual data which identifies the patient included in this study.

### Availability of data and materials

The datasets used and/or analyzed during the current study are available from the corresponding author upon reasonable request.

### Author contributions

BKS designed the study, collected the patient data, and drafted the manuscript.ZJY followed up with patients for overall survival and revised the manuscript. SL assisted in data collection from the central database of the unit.YNZ and ZLZ revised the drafts of the manuscript.CL and ZGZ participated in the design of the study and critically revised the drafts of the manuscript. All authors meet the criteria for publication; all authors have read and approved the final manuscript.

## Acknowledgments

The authors thank all the clinicians in the gastrointestinal department for their support to conduct this study. And especially thanks to Mrs. Qin Yu for recording the data in the central database.

